# Systematic Review of Population-Based Studies of Prevalence and Incidence of Aging-Associated Neurodegenerative Diseases in Russia

**DOI:** 10.64898/2026.04.03.26350047

**Authors:** Okhotin Artemiy, Gorbunova Irina, Bolshakov Anton

## Abstract

**Aim:** To systematically review all population-based studies reporting the prevalence or incidence of neurodegenerative diseases among adults aged 50 and older in Russia.

**Subject and Methods:** We systematically searched Medline, Scopus, Embase, and eLibrary from inception to January 2025. The search included peer-reviewed articles, grey literature, and citation checking. Cross-sectional and cohort studies were eligible if they reported population-based prevalence estimates of dementia, cognitive impairment, or Parkinson’s disease in community-dwelling adults aged 50 years and older in Russia. Studies performed in healthcare settings and insitutionalized populations were excluded. The risk of bias was assessed using the RoB-PrevMH tool. For studies based on cognitive screening tools, dementia prevalence was adjusted for test sensitivity and specificity. Meta-analysis of available prevalence and incidence estimates was performed using random– and fixed-effects models for proportions, with pooling stratified by age group and assessment method.

**Results:** A total of 20 studies met the inclusion criteria. Diagnostic methods varied, including expert evaluation, cognitive screening instruments (e.g., MMSE, Mini-Cog), and administrative data. Prevalence estimates for dementia ranged from 0.5% to 81.6%, with the lowest values derived from administrative data and the highest from Mini-Cog screening among adults aged 85 and older. Cognitive dysfunction was reported in 12 studies, with prevalence estimates ranging from 3.1% (MMSE<24, age 55–74) to 81.5% (Mini-Cog, age 85 and older). Nine studies reported the prevalence of Parkinson’s disease, ranging from 0.017% to 0.29% in administrative data, and 0.31% in the only study using neurologist assessments.

**Conclusion:** The prevalence of dementia and Parkinson’s disease in Russia varies widely depending on diagnostic method, age group, and study design. Most studies lacked representative sampling and used non-standardised diagnostic criteria. These findings underscore the importance of population-based epidemiological research in supporting public health planning and policy development within the Russian context.

## 1 Introduction

Neurodegenerative diseases are an important public health problem. The prevalence and impact increase with population ageing.[1] According to the Global Burden of Disease project, the number of people living with dementia in 2019 is estimated at 57 million and will increase almost threefold to 153 million by 2050.[2] Incidence and prevalence depend on the demographic structure and prevalence of risk factors; some of them, such as education, social isolation, smoking and alcohol use, are economically and culturally determined.[1] Country-specific data on epidemiology are necessary to inform public health policies.

Population of Russia is ageing and there is need for epidemiological studies examining the health and well-being of elderly people.[3] The aim of this work is to systematically review all published studies that report the incidence and prevalence of the most common neurodegenerative diseases in the elderly, specifically Alzheimer’s disease, dementia, mild cognitive impairment, and Parkinson’s disease.

All cohort and cross-sectional studies reporting the incidence and prevalence of these conditions in people 50 years and older in Russia will be reviewed and critically appraised.

## 2 Results

A search in databases resulted in 947 unique articles; 47 additional articles were found after reference lists and website searches. Of them, 947 articles were excluded based on title and abstract screening, and 20 were excluded after full-text screening, leaving a total of 20 eligible articles: 12 papers assessing prevalence and incidence of cognitive disorders and 9 papers estimating prevalence and incidence of Parkinson disease. The process is detailed in Figure 1.

**Figure 1.**
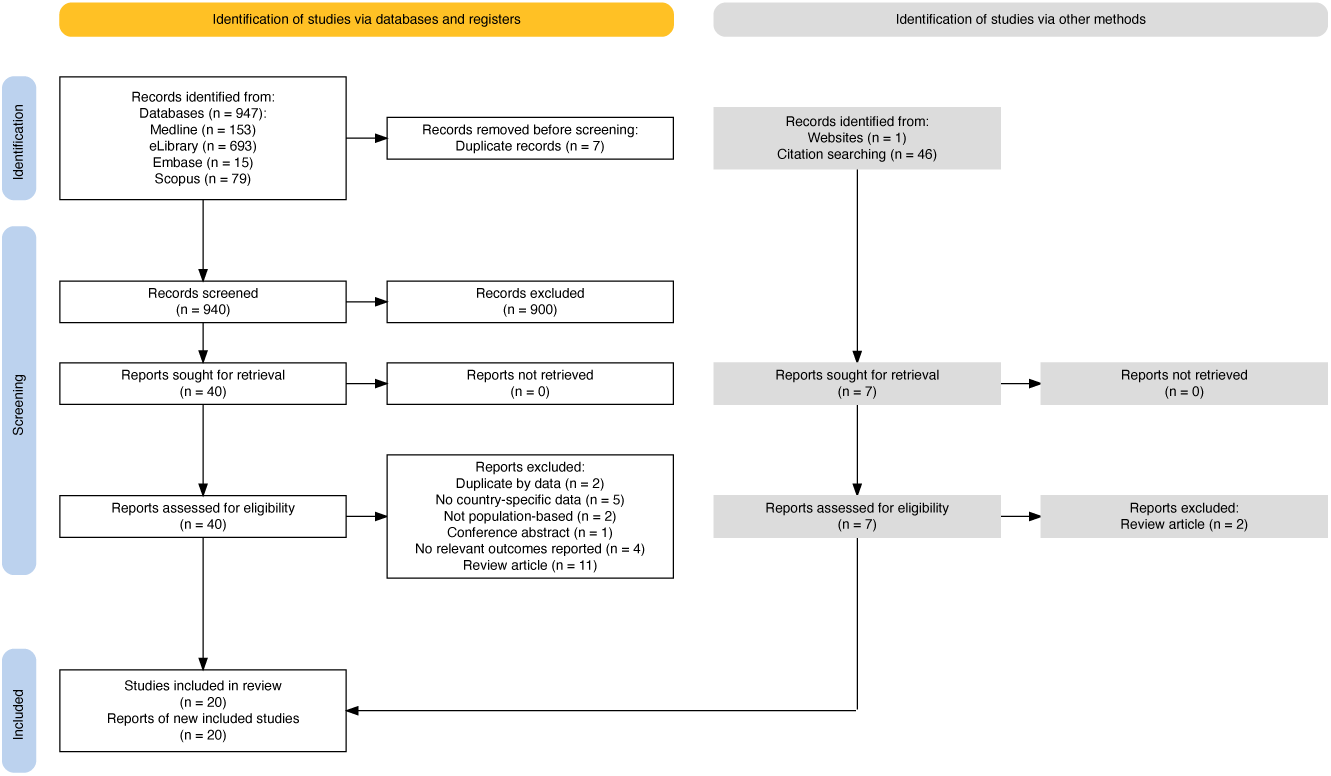
PRISMA Diagram.

### 2.1 Studies of cognitive disorders

There were a total of 12 papers describing studies assessing cognitive outcomes.[4–9, 9–14]. Three of these papers describe one cohort (CRYSTAL study)[6–8], and two describe another cohort (EVKALIPT study)[10, 13], overall comprising a total of 9 study populations.

CRYSTAL study performed in Kolpino district in Saint-Petersburg was reported separately for two age cohorts: 65—74 years and 75 years and older. EVKALIPT study, performed in 11 regions, was reported for three age cohorts (65—74 years, 75—84 years, 85 years and older) and also for different groups of regions separately. In other studies, data were not reported for different age cohorts. Most of the studies were cross-sectional. Two were based on routinely collected healthcare administrative data[9, 11]. Only one cohort study assessed the incidence of cognitive decline[8]; all others were cross-sectional.

In most studies, only cognitive dysfunction, as defined by cognitive test results below the cut-off point, was reported, and dementia prevalence and confidence intervals were projected by us. In some cases, reported prevalences of cognitive dysfunction were incompatible with published sensitivities and specificities of the cognitive tests used. In two studies, dementia was diagnosed by experts.[4, 5] Two studies are based on administrative data.[9, 11]. Prevalence of cognitive dysfunction in the CRYSTAL study was reported separately based on different cut-offs of MMSE score; we provide both estimates. Results on prevalence of dementia and cognitive dysfunction are presented in Table 1.

**Table 1.** Dementia and Cognitive Dysfunction Prevalence Estimates.

| Study | Age | Reported outcome | Diagnostic criteria | Cognitive dysfunction | Dementia |
| --- | --- | --- | --- | --- | --- |
| Moscow, 1977 | older than 60 | Dementia | Psychiatrist | - | 6.7 (5.2–8.3) |
| Moscow, 2001 | 60 and older | Dementia (Alzheimer's, vascular) | Psychiatrist | - | 10.5 (8.7–12.3) |
| Chuvash Republic, 2020 | older than 60 | Dementia | Administrative data | - | 0.61 |
| Northwest, 2021 | older than 60 | Dementia | Administrative data | - | 5.2 |
| CRYSTAL, 2011 | 65 and older | Dementia | MMSE<24 | 26.3 (22.9–29.8) | 21.6 (17.1–26.6) |
| CRYSTAL, 2011 | 65–74 | Dementia | MMSE<24 | 12.9 (9.2–16.7) | 3.3 (0.0–8.9) |
| CRYSTAL, 2011 | 75 and older | Dementia | MMSE<24 | 40 (34.5–45.4) | 39.8 (32.7–47.5) |
| CRYSTAL, 2011 | 65 and older | Cognitive dysfunction | MMSE<25 | 34.6 (30.9–38.4) | 24.0 (18.5–29.8) |
| CRYSTAL, 2011 | 65–74 | Cognitive dysfunction | MMSE<25 | 20.4 (15.9–24.9) | 2.9 (0.0–10.1) |
| CRYSTAL, 2011 | 75 and older | Cognitive dysfunction | MMSE<25 | 49 (43.5–54.6) | 45.0 (36.8–53.5) |
| EVKALIPT Subgroup, 2020 | 65 and older | Cognitive dysfunction | Mini-Cog<=3 | 63.9 (60.3–67.6) | 75.5 (52.5–100.0) |
| EVKALIPT Subgroup, 2020 | 65–74 | Cognitive dysfunction | Mini-Cog<=3 | 45 (38.6–51.3) | 36.5 (19.5–61.4) |
| EVKALIPT Subgroup, 2020 | 75–84 | Cognitive dysfunction | Mini-Cog<=3 | 66.7 (60.5–72.9) | 81.6 (54.9–100.0) |
| EVKALIPT Subgroup, 2020 | 85 and older | Cognitive dysfunction | Mini-Cog<=3 | 81.5 (76.3–86.7) | Incompatible |
| Ural Very Old, 2022 | 85 and older | Cognitive dysfunction | MMSE<24 | 48.6 (46–51.2) | 51.5 (47.8–55.5) |
| EVKALIPT (11 regions), 2022 | 65 and older | Cognitive dysfunction | Mini-Cog<=3 | 60.8 (59.2–62.4) | 69.1 (48.6–100.0) |
| ESSE-RF, 2023 | 55–74 | Cognitive dysfunction | MMSE<24 | 3.1 (2.8–3.4) | Incompatible |

There was only one longitudinal study[8], which assessed incidence of cognitive decline in people of 65 years or older. It reports clinically relevant decline in MMSE score in 41.0% people during 2.5 years (without confidence interval of the estimate).

Most studies had a low risk of bias for sample selection (Criterion 1). Risk for non-response bias (Criterion 2) was more mixed — several studies lacked sufficient reporting or relied on administrative data, leading to a higher risk of uncertainty. The major variability was in Criterion 3 — outcome classification. In studies reporting only cognitive dysfunction based on MMSE and Mini-Cog cognitive scores we assessed this criterion twice: separately for cognitive dysfunction (Figure A3) and for projected dementia prevalence (Figure A4).

Reported estimates are highly heterogeneous and are difficult to compare due to differences in methods, demographics, and populations. Meta-analysis clearly shows that prevalence estimates of dementia vary significantly depending on the age and diagnosis classification (Figure 2). Data on gender-specific prevalences were reported only in two studies, and there was no statistically significant differences between genders.[7, 14]

**Figure 2.**
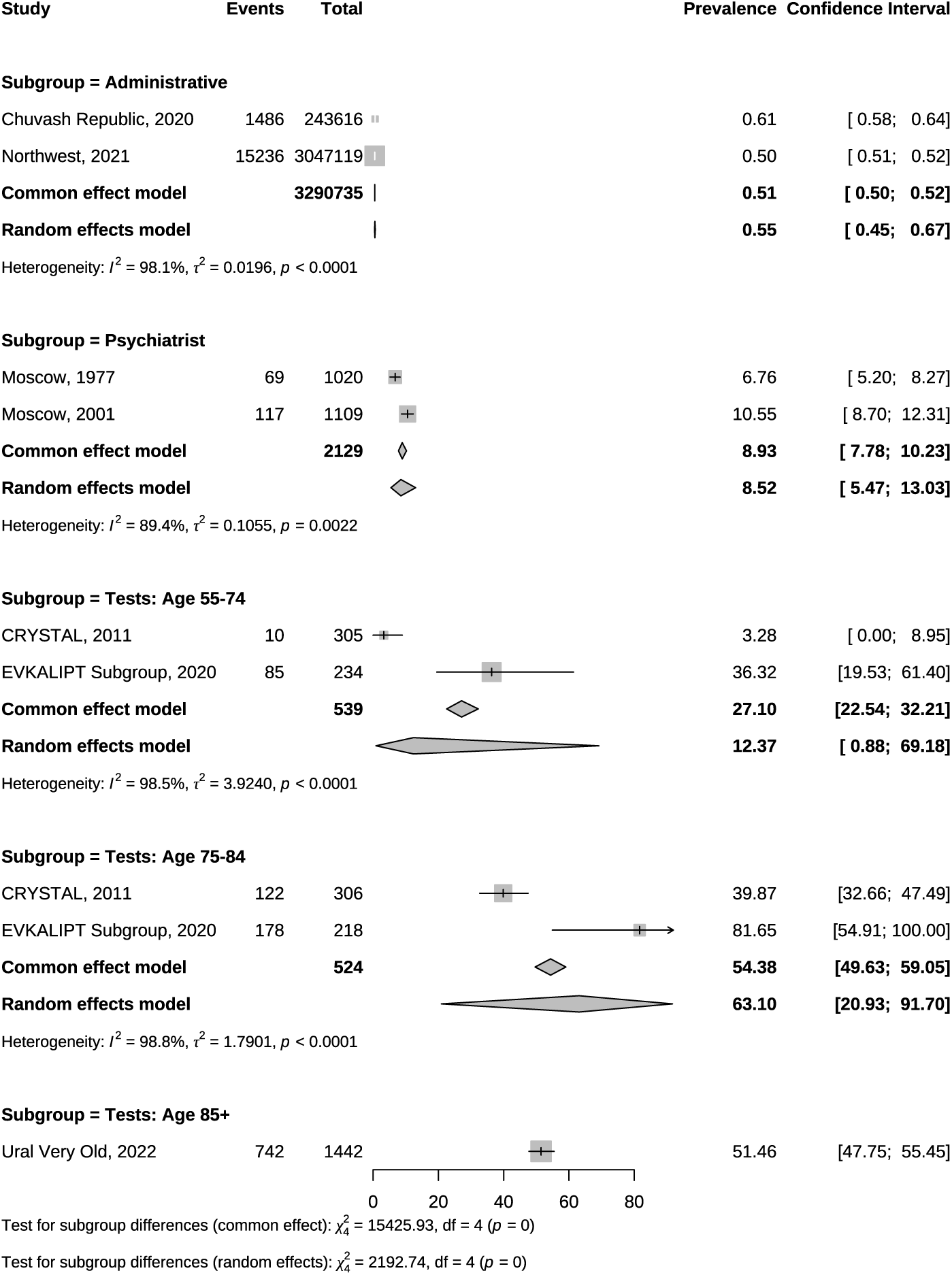
Meta-analysis of Dementia Prevalence Estimates. ^a^ For unadjusted estimates, confidence intervals were calculated using the Agresti–Coull method[24]. ^b^ For studies using cognitive screening tools (MMSE and Mini-Cog), misclassification was accounted for and confidence intervals were calculated using the Lang & Reiczigel method[25].

Projections based on single cognitive test results give the highest estimates of dementia prevalence and show clear increase within age ranges: 12.37% (95% CI: 0.88—69.18) for 55—74 years, 63.1% (95% CI: 20.93—91.70) for 75-84 years and 51.46% (95% CI: 48.84—54.07) for 85 and older (single study). Two studies based on expert diagnosis show the least heterogeneity and mid-range estimated dementia prevalence of 8.52% (95% CI: 5.47-13.07).

Prevalence of dementia as defined by cognitive test score lower than cut-off value is 13.45% (95% CI: 2.40—49.49) for 55—74 years, 53.36% (95% CI: 28.08—77.02) for 75—84 years and 66.99% (95%CI: 30.87-90.22) for 85 years and older. In sensitivity analysis the study with the highest risk of bias due to non-random sampling was excluded, and estimates became lower but still higher than for expert diagnosis-based studies (Figure A2).

### 2.2 Studies of Parkinson’s disease

A total of nine papers were found that describe studies assessing Parkinson’s disease. [15–23]

Most of the reported estimates of the prevalence of Parkinson’s disease (Table 2) are derived from routinely collected healthcare administrative data or data on referral to clinics with known coverage. The only population-based study evaluated all people of 40 years or older in Solnechnogorsk district with population of 4,473 by neurologist with prevalence estimate of 0.31% (95% CI: 0.15–0.47). The remaining studies, relying on administrative data, reported a wide range of prevalence estimates, from as low as 0.022 [23] to as high as 0.29 (Figure 3).[21]

**Figure 3.**
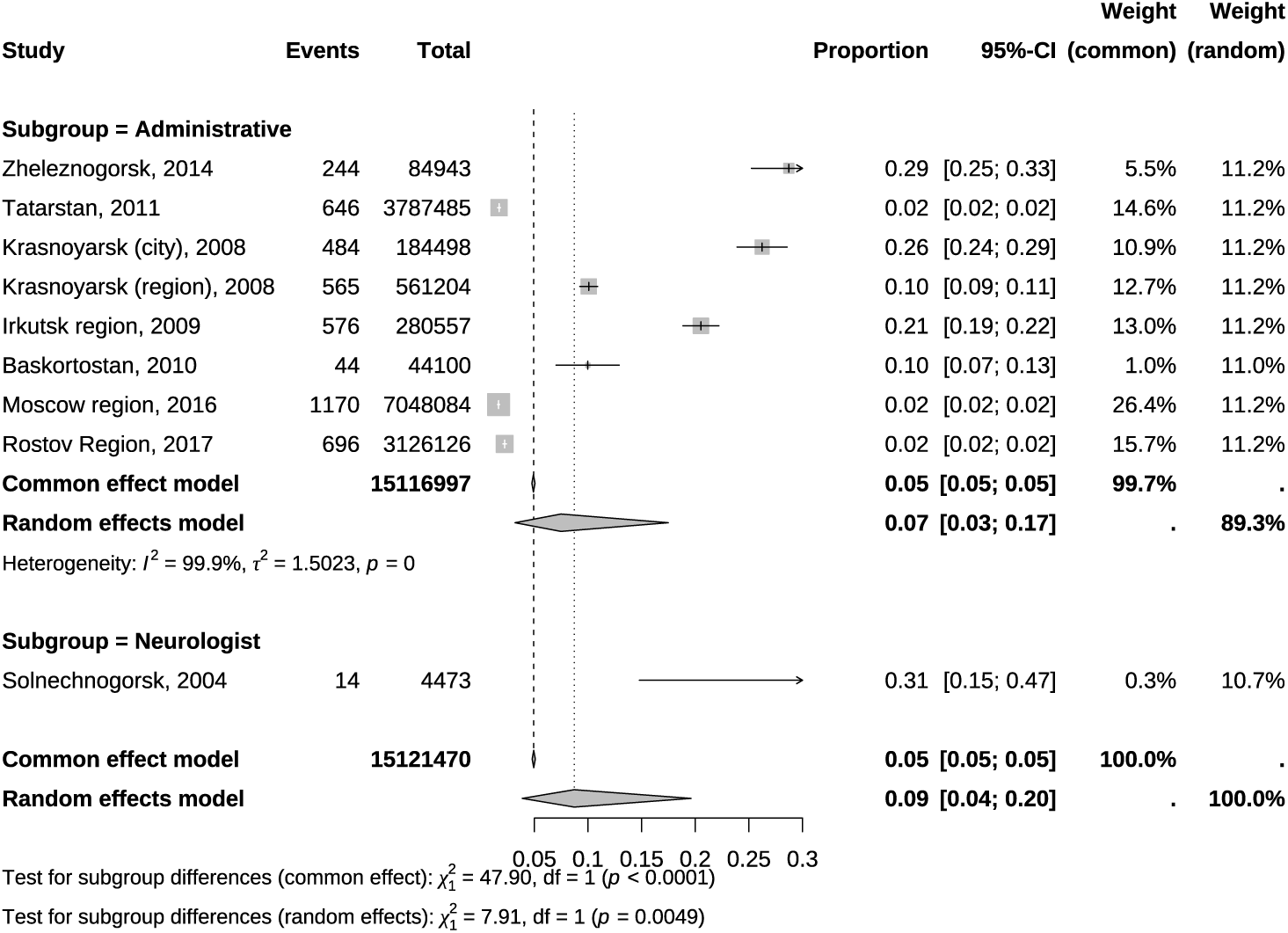
Meta-analysis of Parkinson’s Disease Prevalence Estimates.

**Table 2.** Parkinson’s Disease and Parkinsonism Prevalence and Incidence Estimates.

| Study | Age | Outcomes | Diagnosis | Prevalence, % | Incidence,<br>per 100 000 person-years |
| --- | --- | --- | --- | --- | --- |
| Moscow, 2010 [18] | 60 and older | Parkinson disease | Neurologist | – | 9.03 (8.01–10.15) |
| Solnechnogorsk, 2004 [15] | older than 40 | parkinsonism, Parkinson disease | Neurologist | 0.311 (0.148–0.474) | – |
| Tatarstan, 2011 [20] | all ages | parkinsonism, Parkinson disease | Administrative data | 0.017 (0.016–0.018) | 17.0 |
| Zheleznogorsk, 2014 [21] | older than 40 | Parkinson disease | Administrative data | 0.288 | 13.8 |
| Krasnoyarsk (city), 2008 [16] | older than 65 | Parkinson disease | Administrative data | 0.262 (0.239–0.286) | – |
| Krasnoyarsk (region), 2008 [16] | older than 65 | Parkinson disease | Administrative data | 0.101 (0.092–0.109) | – |
| Irkutsk region, 2007 [17] | older than 65 | Parkinson disease | Administrative data | 0.205 (0.189–0.222) | – |
| Baskortostan, 2010 [19] | all ages | Parkinson disease, parkinsonism | Administrative data | 0.1 (0.07–0.129) | – |
| Moscow region, 2016 [22] | all ages | Parkinson disease, parkinsonism | Administrative data | 0.017 (0.016–0.018) | – |
| Rostov Region, 2017 [23] | all ages | Parkinson disease | Administrative data | 0.022 (0.021–0.024) | – |

Estimates of the incidence of Parkinson’s disease and parkinsonism are presented in three studies [15, 18, 21], with incidence rates for Parkinson’s disease ranging from 9.03 to 17.0 per 100,000 person-years across all age groups. Only one of these studies is population-based[15] while two others are based on referral and administrative data. Due to highly heterogeneous design and population demographics quantitative meta-analysis was not performed.

## 3 Methods

### 3.1 Protocol and registration

This systematic review followed the Preferred Reporting Items for Systematic Reviews and Meta-Analyses (PRISMA) guidelines.[26] The protocol was registered in PROSPERO (CRD42024613249; https://www.crd.york.ac.uk/PROSPERO/view/CRD42024613249).

### 3.2 Eligibility Criteria

We included:

- Observational studies (cohort or cross-sectional) reporting population– or community-based prevalence or incidence of neurodegenerative diseases (e.g., Alzheimer’s disease, dementia, Parkinson’s disease/parkinsonism, cognitive or motor function decline, or other neurodegenerative conditions).
- Studies conducted in Russia or its predecessor in the USSR (Russian Soviet Federative Socialist Republic).
- Participants aged *≥* 50 years.
- Articles published in English or Russian.

We excluded case reports/series, conference abstracts, editorials/commentaries, letters, and reviews without original data. Studies restricted to institutionalised populations (e.g., nursing homes) were excluded due to limited comparability with community-dwelling older adults.

### 3.3 Information Sources and Search Strategy

We searched Medline, Scopus, Embase, and eLibrary (the largest Russian scientific electronic library) from inception to January 25, 2025. Language-specific queries were used in eLibrary to capture Russian-language publications. Search strategies combined terms for observational designs (e.g., cohort, incidence, prevalence), neurodegenerative outcomes (e.g., dementia, Alzheimer’s, Parkinson’s), and geographic terms (Russia/-former USSR), using both controlled vocabulary (e.g., MeSH, EMTREE) and free-text terms. Full strategies are provided in Appendix A.

We also searched Google Scholar and conducted general web searches via Google to identify grey literature, and screened reference lists of included studies and related systematic reviews.

### 3.4 Study selection

Records were screened in two stages. Two reviewers independently screened titles/abstracts, then assessed full texts of potentially eligible studies. Disagreements were resolved with a third reviewer; final decisions were reached by consensus. The most common reasons for exclusion were: not population– or community-based; no country-specific data; and outcomes not relevant to Alzheimer’s disease, dementia, Parkinson’s disease/parkinsonism, cognitive or motor function/decline, or other age-related neurodegenerative diseases.

### 3.5 Data extraction

A standardised, pre-piloted extraction form was used. Three reviewers independently extracted data; discrepancies were resolved by discussion. Data were recorded in Google Sheets. We extracted study design, sample size, participant characteristics (age, gender when available), diagnostic criteria, and cognitive/motor assessment instruments.

Given heterogeneity, outcomes were grouped as cognitive impairment (primarily MMSE and Mini-Cog), all-cause dementia, and Parkinson’s disease. No included studies specifically evaluated Alzheimer’s disease.

For multiple reports from the same cohort, we cross-checked results to avoid duplication and extracted data from the most comprehensive source.

### 3.6 Quality assessment

Two reviewers assessed methodological quality and the principal investigator reviewed ratings using a modified three-item Risk of Bias tool for prevalence studies (RoB-PrevMH).[27] Items covered sampling frame representativeness (sampling bias), representativeness of responders (non-response bias), and information bias. Disagreements were resolved by consensus.

Information bias was rated high when dementia prevalence was based solely on cognitive screening tools due to limited diagnostic accuracy.

For healthcare administrative data or single tertiary care centre datasets serving a defined population, sampling-frame representativeness was rated low risk; non-response bias was rated high because inclusion depended on care-seeking.[28] Information bias was rated high when routine diagnostic criteria were unclear or inconsistent, unless validation or adjustment procedures were reported.

### 3.7 Synthesis of the Results

Because included studies differed substantially in populations and methods, meta-analyses were conducted separately by age group and diagnostic/data source category (administrative data, expert diagnosis, and projected prevalence from single screening tests). Only prevalence was synthesised because incidence was reported in a single study.[18]

For studies using MMSE or Mini-Cog, dementia prevalence was adjusted for test sensitivity and specificity.[29, 30] Confidence intervals for adjusted estimates were calculated using the Lang & Reiczigel method.[25] For administrative-data studies, reported prevalence was summarised without adjustment; confidence intervals for unadjusted estimates used the Agresti–Coull method.[24]

Meta-analyses used metaprop in the *meta* R package (version 8.0.2). Proportions were pooled on the logit scale with inverse-variance weighting; both common-effect and random-effects (REML) models were reported. Subgroup analyses were conducted by diagnostic method (psychiatrist assessment, neurologist assessment, administrative data, screening tests) and age group. Heterogeneity was summarised using *I*^2^ and *τ*^2^, and forest plots presented study-specific and pooled estimates. Sensitivity analyses excluded the study with the highest risk of sampling bias.

## 4 Discussion

This systematic review has found heterogenous studies that can be roughly divided in several groups. The first group is older studies with robust methodology and expert assessment of total population of small geographic areas.[4, 5, 15] Some of these studies are accessible only as doctoral theses and not published in peer reviewed journals, which was not unusual in soviet science. The second group consists of studies based on healthcare administrative data from institutions serving populations of certain geographic areas. Those studies are essentially administrative reports and are published in local Russian language journals.[9, 11, 16, 20–23] The third group is represented by modern epidemiological studies conducted in the rapidly evolving field of gerontology. Those studies are published in Russian and international journals.[6, 10, 12, 13, 18, 19]

The majority of the included studies used a cross-sectional design. Only one was longitudinal, with clearly described methods, and it assessed changes in global cognitive function over time.[8]

Despite very heterogeneous results and variable risks of bias in the studies, there are some obvious trends. First, prevalence of both cognitive and motor disorders increases with age. This is in line with international data. Second, prevalence is lowest in administrative data, highest in studies based on cognitive tests, and midrange in older studies based on expert diagnosis. Third, there is a shift in study focus and design from older studies to newer ones, transitioning from medical diagnosis to a broader context of gerontology. While older studies were based on the total assessment of small geographic areas and focused on medical diagnosis [5, 15, 31], they lacked data on the social context of the disease and relied on expert diagnoses without internationally validated tests, which limits comparability with international data. Still, those studies provided the most rigorous estimates of the prevalence of dementia and Parkinson’s disease.

In more recent studies cognitive dysfunction was assessed with internationally validated tests and was studied in the context of other aging-related issues, such as frailty and comorbidity, but still lacked a socio-economic context of disease — care burden and the cost of the disease. Also, dementia was not the primary concern of those studies; its prevalence was projected by us based on the results of a single cognitive tests. Those studies represent significant progress in the field of gerontology in Russia in recent years.

Healthcare administrative data based studies provide estimates without clear context, resulting in poor comparability with other types of studies. Estimates from administrative data were significantly lower, possibly related to underdiagnosis.

We believe that older studies based on expert diagnosis and total assessment provide the most sound estimates, which are also in line with data from high– and middle income countries in Asia and Europe. The reported prevalence of dementia at 6.7–10.5% in people older than 60 years is close to estimates of 7.1% in China[32], 5.8% in Brazil[33], and 10% to 11% in people aged 65 years or older in Europe[34] and the United States[35]. The prevalence of Parkinson’s disease in an older study with total population assessment is 162 cases per 100,000 people [15], which is also consistent with global data on the age-standardized prevalence estimate of 129 cases per 100,000 people [36].

Cognitive dysfunction prevalence in people aged 65 years and older, as assessed by a single cognitive score (26.3–63.9%), is higher in reported studies compared to international estimates of 22% in the United States[35], 8.1% in Brazil[33], and is close to 41% in China[37]. Probably, the prevalence of non-dementia cognitive dysfunction is overestimated in these studies due to the low specificity of MMSE and Mini-Cog scores, as well as the overrepresentation of hospitalized participants in study with the highest estimates.

Underdiagnosis of dementia and Parkinson’s disease is reported across all health-care systems, which means that patients and their families have limited access to specialized medical and social care. The underdiagnosis rate in Russia can be roughly projected from our data: comparing dementia prevalence estimates of 6.7–10.5% in studies based on expert diagnosis with the administrative data estimates of a 0.5–0.6% provides underdiagnosis rate of over 90%. It is close to 90% in India and 95% in Thailand, but is higher than 70% in Brazil and 50% in Europe.[38–41]

Using the most robust population-based estimate of Parkinson’s disease prevalence of 0.162% and administrative estimates of only 0.017–0.02%, administrative data appear to miss around 90% of Parkinson’s disease cases — a degree of underdiagnosis similar to that observed for dementia. While this high rate of underdiagnosis might be overestimated due to poor registration of cases, it is still likely to be very high.

The risk of bias was lowest in the older studies that assessed the total population, which had a high response rate and expert diagnostic classification. While the diagnosis was not based on standardized criteria, it was standard practice at that time. Administrative data based studies were considered to have a high risk of bias, both due to the low response rate (which depended on referral) and due to non-standardized diagnostic classification in real practice. More recent studies had a low risk of bias due to sampling by design; however, sampling was not always performed according to the protocol. Classification bias risk was not inherent to the studies but was introduced due to the projection of dementia prevalence based on global cognitive scores with non-perfect sensitivity and specificity. Overall, the least biased estimates were reported in older studies with a total assessment of small geographical areas; however, they do not represent larger populations.

Thus, our systematic review shows that older studies have robust expert-based assessments of the outcomes but lack data on the context of the disease and represent very small populations. More recent large-scale studies were more representative, but outcome assessment with a single test lacks sensitivity and specificity. Administrative reports provide data on the registration of the disease but have not provided reliable estimates of the true prevalence. There were no studies with robust estimates of incidence.

We believe that the epidemiology of cognitive and motor diseases in the elderly should be studied longitudinally and in the context of aging studies, assessing both medical and social impact of the disease, such as SHARE and HRS family studies.[42, 43] While such studies have been conducted in Russia, they were primarily focused on socioeconomic issues without attention to medical conditions. Any disease is a product of biological processes and social interactions, but this is especially true for cognitive and motor disorders of the older age. So, we assume that any epidemiological study of these diseases should evaluate not only the diagnosis but also its impact on the personal, social, and economic well-being of both individuals and their families and caregivers. These data should become the foundation for evidence-based policies regarding the elderly and the ageing population.

## 5 Conclusion

We identified 20 population-based studies: 12 on cognitive outcomes and 8 on Parkinson’s disease. However, only six represented distinct non-administrative population-based cohorts for cognitive outcomes, and only one for Parkinson’s disease; the remaining evidence largely comes from administrative sources and/or multiple reports from the same cohorts.

Dementia prevalence ranged from 0.5% to 81.6%, depending on age and method. Parkinson’s disease prevalence ranged from 0.017% to 0.311%, mostly based on administrative data. Only one longitudinal study reported the incidence of cognitive decline, and three reported the incidence of Parkinson’s disease. Other studies were cross-sectional, covered selected regions of the country, and did not use standardized diagnostic criteria. Reliable national and regional estimates of prevalence and incidence are lacking. Longitudinal studies with validated tools that assess not only medical diagnoses but also related socioeconomic variables are needed to guide policies.

## Availability of data, code, and other materials

The protocol is registered in PROSPERO (CRD42024613249).

We make the following materials publicly available upon publication:

(i) fully reproducible analytic code (R) used for data cleaning, misclassification adjustment, meta-analyses, forest and traffic-light plots, and automated construction of the PRISMA flow diagram;
(ii) machine-readable tables of all records retrieved with screening decisions and reasons for exclusion;
(iii) the complete data-extraction table with all study-level fields and the aggregated counts and denominators used in syntheses;

The datasets and the R code for the analysis can be accessed directly via the following OSF link: https://doi.org/10.17605/OSF.IO/4BPTV

## 8 Author Contributions

All authors contributed to the study conception and design. Literature searching was performed by all authors. Study selection (eligibility assessment) and risk of bias assessment were performed independently by Irina Gorbunova and Anton Bolshakov; discrepancies were resolved through discussion with Artemiy Okhotin. Data extraction was performed by all authors. Data synthesis was conducted by the study team; figures and meta-analysis were conducted by Irina Gorbunova. The first draft of the manuscript was written by Artemiy Okhotin, and all authors commented on previous versions. All authors read and approved the final manuscript.

## 9 Ethics approval

Not applicable. This study is a systematic review of published and publicly available literature and did not involve recruitment of human participants, use of identifiable individual-level data, or animal experiments; therefore, ethics approval was not required.

## 10 Consent to participate

Not applicable.

## 11 Consent to publish

Not applicable.

## Data Availability

We make the following materials publicly available upon publication:
(i) fully reproducible analytic code (R) used for data cleaning, misclassification adjustment, meta-analyses, forest and traffic-light plots, and automated construction of the PRISMA flow diagram;
(ii) machine-readable tables of all records retrieved with screening decisions and reasons for exclusion;
(iii) the complete data-extraction table with all study-level fields and the aggregated counts and denominators used in syntheses;
The datasets and the R code for the analysis can be accessed directly via the following OSF link: https://doi.org/10.17605/OSF.IO/4BPTV

## Appendix A Figures

**Figure A1.**
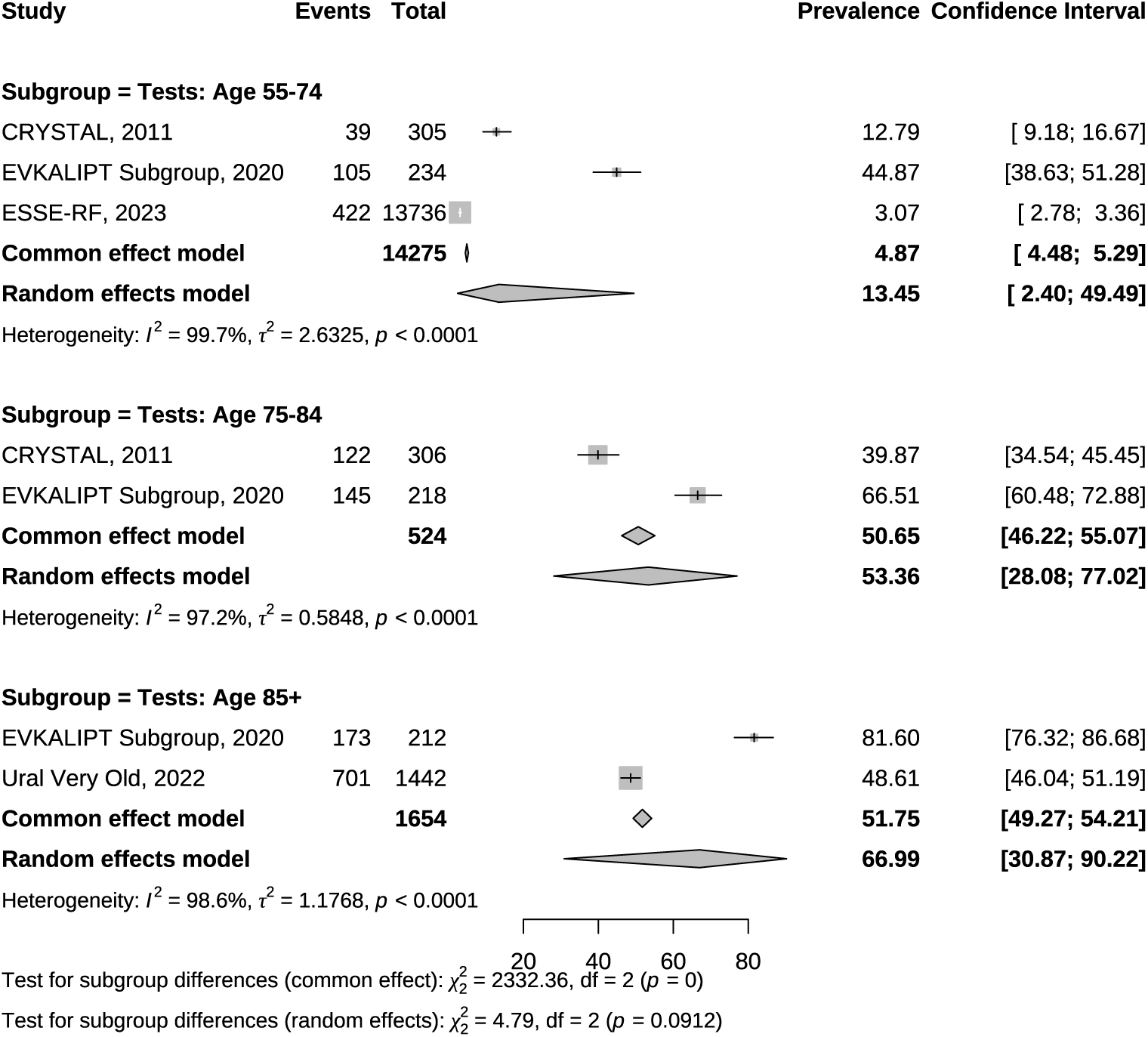
Meta-analysis of Cognitive Dysfunction Prevalence Estimates.

**Figure A2.**
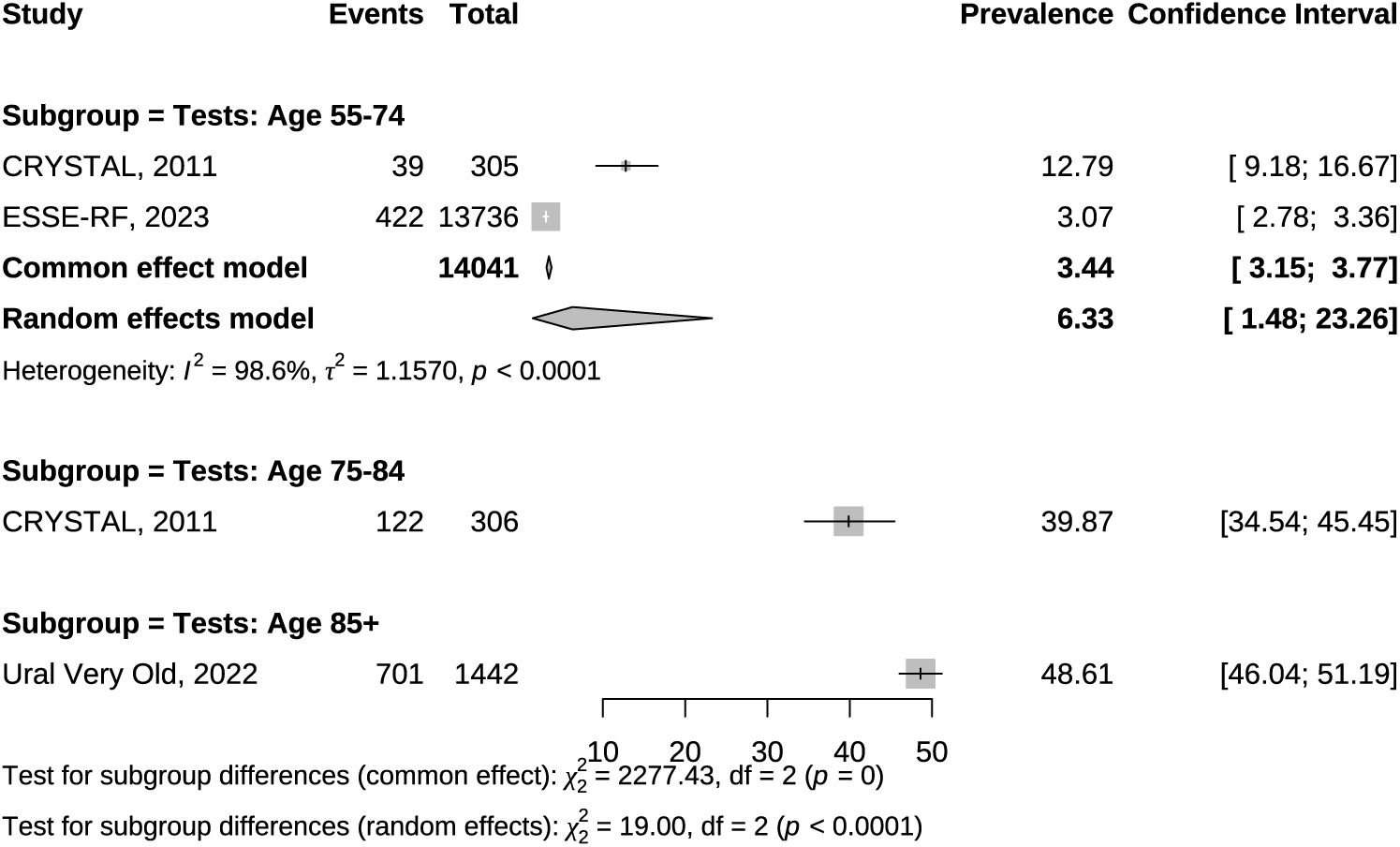
Meta-analysis of Cognitive Dysfunction Prevalence Estimates without Study with High Risk of Sampling Bias[10].

**Figure A3.**
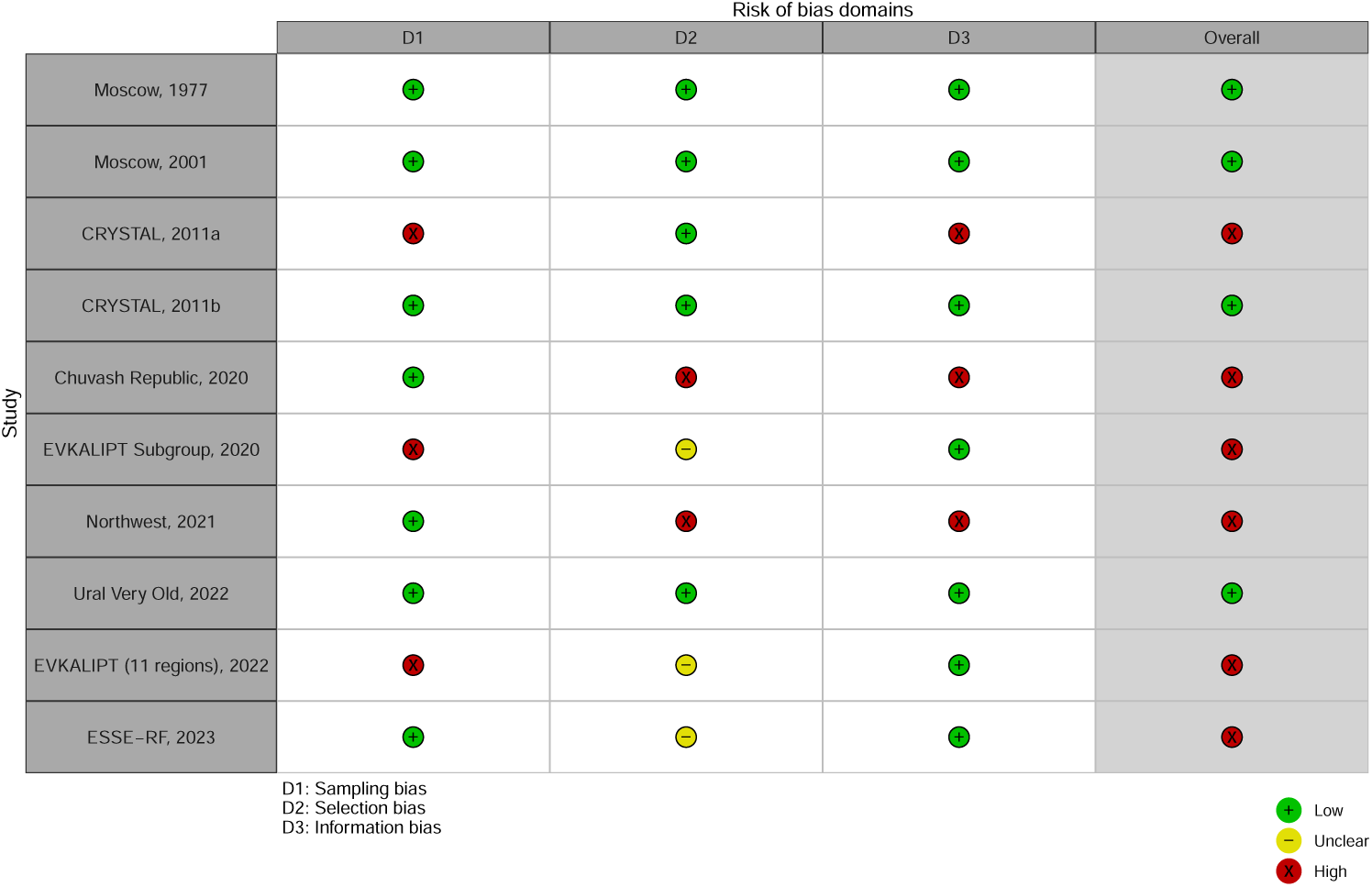
Risk of Bias of Cognitive Dysfunction Assessment. ^a,b^ MMSE cut-offs differed across studies: a, <24; b, <25.

**Figure A4.**
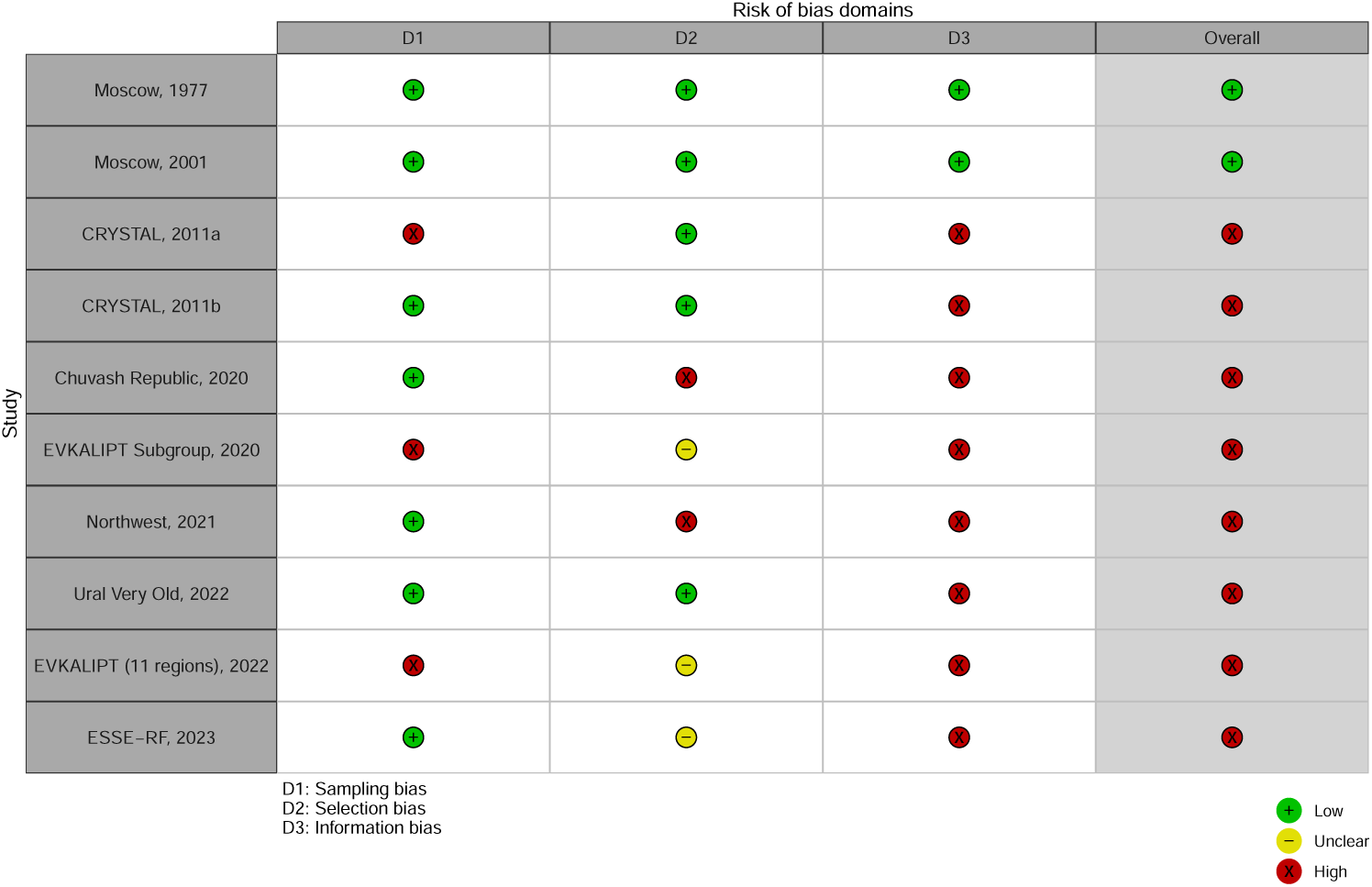
Risk of Bias of Dementia Assessment.

**Figure A5.**
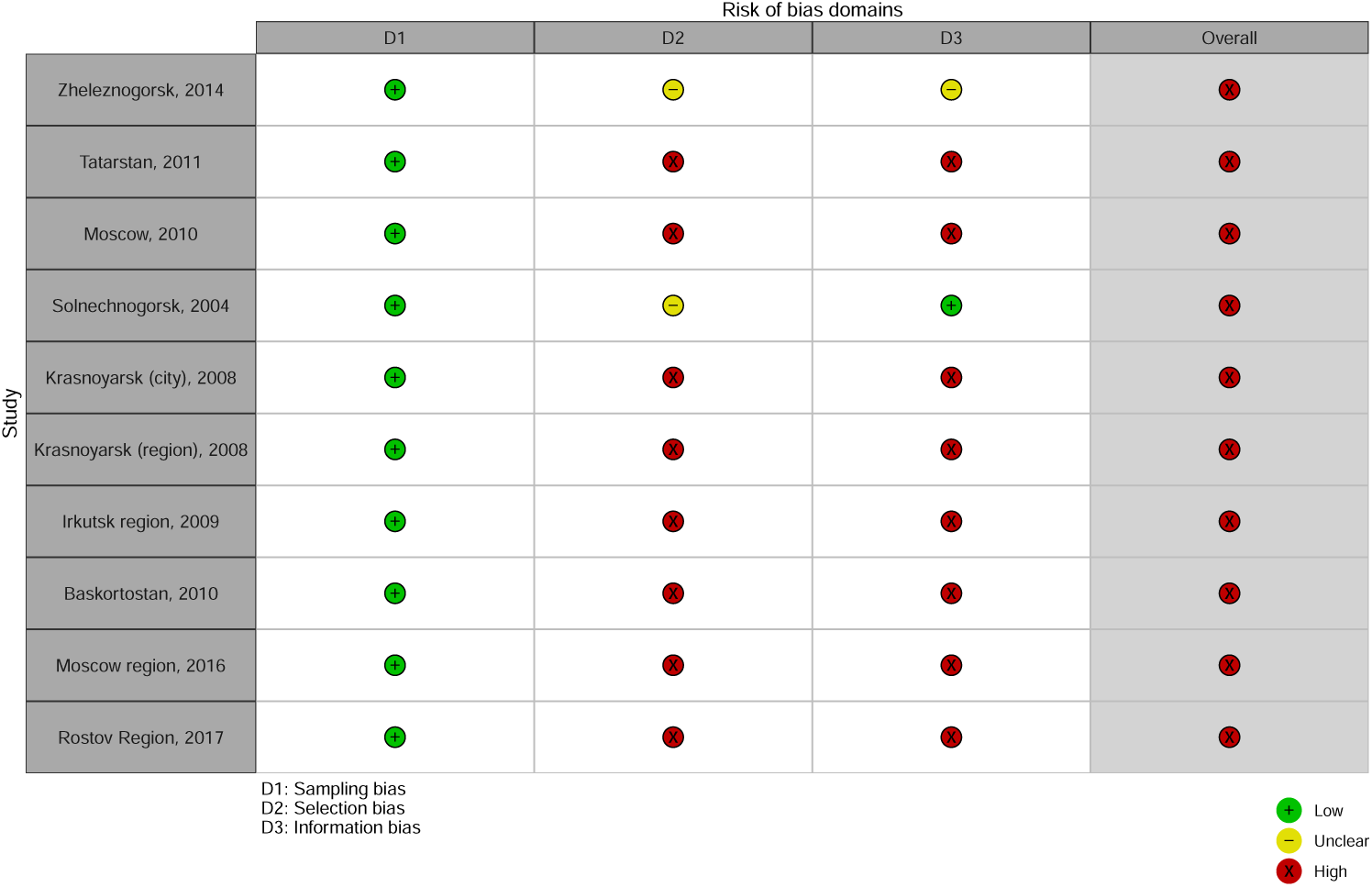
Risk of Bias of Parkinson’s Disease Assessment.

**Table A1.** Cognitive disorders studies.

| Cohort | Location | Age | Population | Outcomes |
| --- | --- | --- | --- | --- |
| Moscow, 1977[4] | Moscow | older than 60 | 1020 | dementia, mild mental deterioration |
| Moscow, 2001[5] | Moscow | 60 and older | 1109 | Dementia (Alzheimer's, vascular) |
| CRYSTAL, 2011[6, 7] | St. Petersburg | 65 and older | 611 | MCI, dementia (mild, moderate, severe) |
| CRYSTAL, 2017 [8]<sup>b</sup> | St. Petersburg | 65 and older | 379 | cognitive decline |
| Chuvash Republic, 2020[9] | Chuvash Republic | older than 60 | 243616 | vascular, Alzheimer dementia |
| EVKALIPT Subgroup, 2020[10]<sup>c</sup> | Moscow, Voronezh | 65 and older | 664 | cognitive dysfunction |
| Northwest, 2021 [11] | Northwestern Federal District | older than 60 | 304712 | vascular dementia |
| Ural Very Old, 2022 [12] | Bashkortostan | 85 and older | 1442 | cognitive dysfunction |
| EVKALIPT (11 regions), 2022 [13] | 11 regions | 65 and older | 3545 | cognitive dysfunction |
| ESSE-RF, 2023 [14] | 14 regions | 55-74 | 13736 | cognitive dysfunction |

<sup>a</sup> Subcohort of CRYSTAL, 2011.<sup>b</sup> Subcohort of EVKALIPT (11regions), 2022.

**Table A2.**
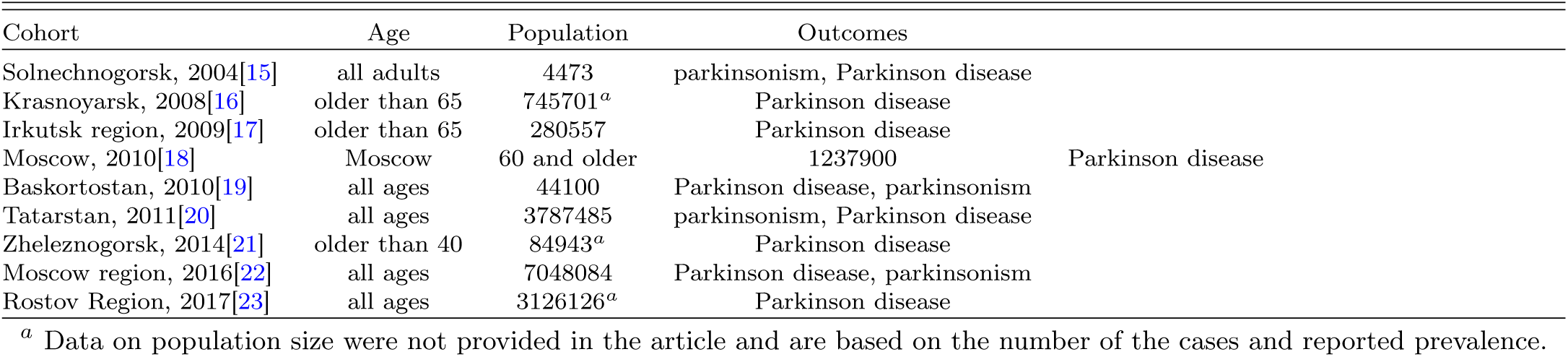
Parkinson’s disease cohorts.

## Appendix B Search Strategy

### Medline

(“cohort studies”[MeSH Terms] OR “cohort study”[Title/Abstract] OR “incidence”[MeSH Terms] OR “incidence”[Title/Abstract] OR “longitudinal studies”[MeSH Terms] OR “longitudinal study”[Title/Abstract] OR “representative group”[Title/Abstract] OR “observational studies”[Title/Abstract] OR “prevalence”[MeSH Terms] OR “prevalence”[Title/Abstract]) AND (“alzheimer disease”[MeSH Terms] OR “alzheimer disease”[Title/Abstract] OR “brain aging”[Title/Abstract] OR “cognitive decline”[Title/Abstract] OR “cognitive dysfunction”[Title/Abstract] OR “dementia”[MeSH Terms] OR “dementia”[Title/Abstract] OR “Dementia/epidemiology”[MeSH] OR “mild cognitive impairment”[Title/Abstract] OR “neurodegenerative disease”[Title/Abstract] OR “neurodegenerative diseases”[MeSH Terms] OR “parkinson disease”[MeSH Terms] OR “parkinson disease”[Title/Abstract] OR “parkinsonism”[Title/Abstract] OR “cognitive impairment”[Title/Abstract] OR “cognitive dysfunction”[MeSH Terms] OR “parkinsonian disorders”[MeSH Terms]) AND (“russia”[MeSH Terms] OR “russia”[Title/Abstract] OR “Russia/epidemiology”[MeSH] OR “russian federation”[Title/Abstract] OR “USSR”[MeSH] OR “USSR”[Title/Abstract])

### Scopus

(INDEXTERMS(“cohort studies”) OR TITLE-ABS-KEY(“cohort study”) OR INDEXTERMS(“incidence”) OR TITLE-ABS-KEY(“incidence”) OR INDEXTERMS(“longitudinal studies”) OR TITLE-ABS-KEY(“longitudinal study”) OR TITLE-ABS-KEY(“representative group”) OR TITLE-ABS-KEY(“observational studies”) OR INDEXTERMS(“prevalence”) OR TITLE-ABS-KEY(“prevalence”)) AND (INDEXTERMS(“alzheimer disease”) OR TITLE-ABS-KEY(“alzheimer disease”) OR TITLE-ABS-KEY(“brain aging”) OR TITLE-ABS-KEY(“cognitive decline”) OR TITLE-ABS-KEY(“cognitive dysfunction”) OR INDEXTERMS(“dementia”) OR TITLE-ABS-KEY(“dementia”) OR INDEXTERMS(“Dementia/epidemiology”) OR TITLE-ABS-KEY(“mild cognitive impairment”) OR TITLE-ABS-KEY(“neurodegenerative disease”) OR INDEXTERMS(“neurodegenerative diseases”) OR INDEXTERMS(“parkinson disease”) OR TITLE-ABS-KEY(“parkinson disease”) OR TITLE-ABS-KEY(“parkinsonism”) OR TITLE-ABS-KEY(“cognitive impairment”) OR INDEXTERMS(“cognitive dysfunction”) OR INDEXTERMS(“parkinsonian disorders”)) AND (INDEXTERMS(“russia”) OR TITLE-ABS-KEY(“russia”) OR INDEXTERMS(“Russia/epidemiology”) OR TITLE-ABS-KEY(“russian federation”) OR INDEXTERMS(“USSR”) OR TITLE-ABS-KEY(“USSR”))

### Elibrary

((заболеваемость) or (когортное исследование) or (популяционное исследование) or (распространенность) or (когорта) or (эпидемиология) or (эпидемиологическое исследование)) & ((болезнь альцгеймера) or (альцгеймер) or (болезнь паркинсона) or (деменция) or (когнитивная дисфункция) or (когнитивная функция) or (когнитивные нарушения) or (нейродегенеративные заболевания) or (паркинсонизм) or (старение мозга) or (синдром умеренных когнитивных нарушений))

### Embase

(TI=(“cohort studies” OR “cohort study” OR “incidence” OR “longitudinal studies” OR “longitudinal study” OR “representative group” OR “observational studies” OR “prevalence”) OR AB=(“cohort studies” OR “cohort study” OR “incidence” OR “longitudinal studies” OR “longitudinal study” OR “representative group” OR “observational studies” OR “prevalence”) OR AK=(“cohort studies” OR “cohort study” OR “incidence” OR “longitudinal studies” OR “longitudinal study” OR “representative group” OR “observational studies” OR “prevalence”) OR KP=(“cohort studies” OR “cohort study” OR “incidence” OR “longitudinal studies” OR “longitudinal study” OR “representative group” OR “observational studies” OR “prevalence”)) AND (TI=(“alzheimer disease” OR “brain aging” OR “cognitive decline” OR “cognitive dysfunction” OR “dementia” OR “dementia/epidemiology” OR “mild cognitive impairment” OR “neurodegenerative disease” OR “neurodegenerative diseases” OR “parkinson disease” OR “parkinsonism” OR “cognitive impairment” OR “cognitive dysfunction” OR “parkinsonian disorders”) OR AB=(“alzheimer disease” OR “brain aging” OR “cognitive decline” OR “cognitive dysfunction” OR “dementia” OR “dementia/epidemiology” OR “mild cognitive impairment” OR “neurodegenerative disease” OR “neurodegenerative diseases” OR “parkinson disease” OR “parkinsonism” OR “cognitive impairment” OR “cognitive dysfunction” OR “parkinsonian disorders”) OR AK=(“alzheimer disease” OR “brain aging” OR “cognitive decline” OR “cognitive dysfunction” OR “dementia” OR “dementia/epidemiology” OR “mild cognitive impairment” OR “neurodegenerative disease” OR “neurodegenerative diseases” OR “parkinson disease” OR “parkinsonism” OR “cognitive impairment” OR “cognitive dysfunction” OR “parkinsonian disorders”) OR KP=(“alzheimer disease” OR “brain aging” OR “cognitive decline” OR “cognitive dysfunction” OR “dementia” OR “dementia/epidemiology” OR “mild cognitive impairment” OR “neurodegenerative disease” OR “neurodegenerative diseases” OR “parkinson disease” OR “parkinsonism” OR “cognitive impairment” OR “cognitive dysfunction” OR “parkinsonian disorders”)) AND (TI=(“russia” OR “russia/epidemiology” OR “russian federation” OR “ussr”) OR AB=(“russia” OR “russia/epidemiology” OR “russian federation” OR “ussr”) OR AK=(“russia” OR “russia/epidemiology” OR “russian federation” OR “ussr”) OR KP=(“russia” OR “russia/epidemiology” OR “russian federation” OR “ussr”))

